# Overweight-Obesity And Glucose Intolerance In Offspring Of Indian Diabetic Mothers

**DOI:** 10.1101/2021.05.17.21257222

**Authors:** Sonali S. Wagle, Sanat Phatak, Shubha Ambardekar, Bhat Dattatrey, Madhura K. Deshmukh, Rajashree Kamat, Sayali Wadke, Shivani Rangnekar, Rasika Ladkat, Kalyanaraman Kumaran, Pallavi C. Yajnik, Chittaranjan S. Yajnik

## Abstract

**Aims:** Maternal diabetes in pregnancy increases offspring obesity and diabetes risk. We investigated body size and composition, and glucose tolerance in offspring born to Indian diabetic mothers (ODM) and to non-diabetic mothers (ONDM), and studied maternal and paternal determinants.

**Methods:** We compared the physical characteristics, body composition (Dual energy X-ray Absorptiometry) and glycemia of ODMs and matched ONDMs. Overweight-obesity was defined using International Obesity Task Force (IOTF) for 2-18 years (cutoff of BMI > 25 kg/m^2^) and World Health Oraganization (WHO) criteria for >18 years (BMI > 25 kg/m^2^). Glycemic measures included capillary blood glucose measurement in children <10 years of age and a 1.75g/kg glucose OGTT in those >=10 years. We calculated separate SD scores for capillary fasting, capillary random and venous fasting plasma glucose. Those above median SD score were classified as glucose intolerant. We evaluated insulin sensitivity (Homeostatic Model Assessment HOMA-S and Matsuda index), beta cell function (HOMA-β and insulinogenic index) and β-cell compensatory response (Disposition Index: [Log (Insulinogenic index) + Log (Matsuda index)]). We studied the association of maternal and paternal body size and glycemia with outcomes in the child.

**Results:** We studied 200 ODMs of 176 diabetic mothers (133 GDM, 21 type 2 diabetes, 22 type 1 diabetes), and 177 ONDMs at an average of 9.7 years after delivery. ODMs were heavier, more adipose and more glucose intolerant than ONDMs. Differences for body size parameters were more prominent in males and they also had a wider spectrum of metabolic abnormalities. Three (4%) ODM were receiving treatment for diabetes (diagnosed between 10-25 years of age). On OGTT, the older ODMs (>= 10 years) had higher prevalence of glucose intolerance (1 DM, 14 IFG, 12 IGT and 4 both IFG and IGT) compared to ONDM, (0 DM, 7 IFG, 9 IGT and 1 both IFG and IGT). None of the diabetic and pre-diabetic ODMs, including children of type 1 diabetic mothers, were positive for circulating GAD or ZnT8 antibodies.

Younger ODMs (<10 years) also had higher capillary blood glucose concentrations compared to ONDM. Overall, ODMs had higher prevalence of glucose intolerance compared to ONDMs, both in younger and older, and in boys and girls. HOMA-S and Disposition index were lower in ODM compared to ONDM. Other indices of insulin secretion and action (HOMA-β, Insulinogenic index and Matsuda index) were similar in the two groups.

Type 2 diabetic and GDM mothers were heavier compared to type 1 diabetic mothers, and their children were more likely to be overweight-obese. Children of type 1 diabetic mothers were glucose intolerant despite lack of overweight-obesity. In addition, fathers had an independent influence on the child’s phenotype, especially for overweight-obesity. Maternal hyperglycemia during pregnancy had an overriding influence on offspring glucose intolerance.

**Conclusions:** ODMs were more overweight-obese and glucose intolerant compared to ONDMs. We propose that these two outcomes in the ODMs are independently programmed by respective parental phenotypes. Preventive strategies will need to be informed by these findings. Studies of genetic and epigenetic mechanisms involved in fetal programming of body size and glycemia will further help our understanding.

**Research in Context:** *What is already known about this subject?:* India has experienced a rapid escalation of diabetes in young individuals including diabetes in pregnancy. Short-term effects of maternal hyperglycemia on the offspring are well known.

*What is the key question?:* There is little data on long-term effects of maternal hyperglycemia on offspring body size and cardiometabolic risk factors. We compared these in the offspring of diabetic mothers compared to those of non-diabetic mothers. We also sought differences within types of diabetes (type 1, type 2, GDM) and studied paternal determinants of these outcomes.

*What are the new findings?:* Type 1 diabetic mothers were thinnest and most hyperglycemic; type 2 diabetic mothers were most overweight-obese, GDM mothers were intermediate. Gestational maternal hyperglycemia was the overriding determinant of offspring hyperglycemia. Maternal hyperglycemia predicted offspring glucose intolerance but not overweight obesity; maternal overweight-obesity predicted offspring overweight-obesity but not hyperglycemia, suggesting an uncoupling of these phenotypes often considered congruent. Fathers had an additive influence on offspring size.

*How might this impact on clinical practice in the foreseeable future?:* Knowing the relative independence of influences on body size and metabolic outcomes will inform strategies of their primordial and primary prevention. Establishing genetic and epigenetic mechanisms will help.

## Introduction

In 2019, an estimated 16% of live births worldwide (>20 million) were exposed to some form of glucose intolerance in pregnancy ^[1]^ including type 1, type 2 diabetes and gestational diabetes mellitus (GDM, glucose intolerance first recognized during pregnancy) ^[2]^. Maternal diabetes affects pregnancy outcomes adversely and possibly contributes to a cascading epidemic of obesity and diabetes in the offspring ^[3–6]^. Both genetic and non-genetic mechanisms (intrauterine programming) are likely contributors, and studies in Pima Indians suggest that the influence of the intrauterine environment may outweigh genetic influences ^[3–5]^. Obesity and glucose intolerance are strongly related in mothers from the western world, making it difficult to distinguish between these two related yet distinct, exposures. Recent reports suggest independent effects of obesity and glucose intolerance, without conclusive evidence. ^[6–10]^ Clarity may come from non-obese diabetic mothers from populations with a lower BMI, such as those in India or those with type 1 diabetes. Analysing paternal influences would further improve our understanding of these complex inter-generational connections.

GDM is common in India, affecting 10 - 15% of pregnancies, and Indian mothers with GDM are younger and have a lower BMI than Europeans ^[11]^. In a previous small Indian follow-up study, 45 offspring of mothers with GDM had higher adiposity and glucose intolerance than controls. ^[12, 13]^ This study did not include forms of diabetes other than GDM.

Our specialized Diabetes Unit has treated hundreds of diabetic pregnancies, and we were able to follow the offspring born to mothers with different types of diabetes and matched controls born to non-diabetic pregnancies. In addition we studied contribution of fathers to the intergenerational associations. We were thus able to investigate independent effects of maternal and paternal obesity and glucose intolerance on the corresponding phenotypes in the offspring.

## Methods

### Study participants

As part of the InDiaGDM (**In**tergenerational programming of **Dia**besity in offspring of women with **G**estational **D**iabetes **M**ellitus) study, we reviewed records of ∼1000 diabetic pregnancies (type 1 diabetes, type 2 diabetes and GDM) treated in our department over last 30 years. We attempted to contact these mothers by telephone or by postal mail to request participation of their offspring (Offspring of Diabetic Mothers, ODM) in the study. Those who agreed to participate were requested to invite their friends, classmates or neighbours as controls if their mothers were not diagnosed with diabetes in pregnancy (Offspring of Non-Diabetic Mothers, ONDM). ONDM were matched for age (±1 year), socioeconomic status and sex with ODMs. We studied child, mother and father from the participating families.

Based on parents’ unwillingness for a venepuncture and full OGTT for young children, we decided to measure a single capillary blood glucose for those below 10 years of age. For those ≥10 years of age a full OGTT was planned (venous sample collected through an indwelling plastic cannula). For the OGTT, participants were advised to follow normal lifestyle for at least 3 days prior to the test. They reported to the Diabetes Unit after an overnight fast (water permitted).

We recorded medical history in the children and parents. Socioeconomic status (SES) of the family was evaluated using the Standard of Living Index (SLI), a tool developed for the National Family Health Survey of India ^[14]^, higher score denotes higher SES.

### Anthropometric measurements

A trained observer made all measurements using standardised protocols ^[15]^. Following measurements were made: weight (Electronic weighing scales, ATCO Healthcare Ltd, Mumbai, India); standing height (wall mounted Harpenden stadiometer, CMS Instruments Ltd, London, UK); circumferences (waist and hip, non-stretchable fiberglass tape, CMS Instruments, London, UK); skinfold thicknesses (bicep, tricep, subscapular and suprailiac, Harpenden skinfold calipers, CMS Instruments, London, UK). An average of two readings was used for analysis. Pubertal staging (Tanner) was done for those between 8 to 18 years of age by a medical officer ^[16]^. We recorded history of menarche in female children.

Overweight-obesity was defined using International Obesity Task Force (IOTF) method for 2-18 years (cutoff of BMI > 25 kg/m^2^) ^[17]^ and World Health Oraganization (WHO) criteria for those >18 years (BMI > 25 kg/m^2^) ^[18]^.

### Body composition

Dual Energy X-ray Absorptiometry (DXA) scans were performed on offspring in pediatric / adult mode, as appropriate, using Lunar Prodigy fan beam machine (GE Medical Systems, Madison, WI, USA) to measure total bone, fat and lean mass.

### Metabolic measurements

In children ≥10 years, an oral glucose tolerance test (OGTT) was performed in fasting state, using 1.75 g of anhydrous glucose per kg body weight up to a maximum of 75 g; post glucose 30 min and 120 min venous samples were collected. For parents we checked fasting capillary glucose by a calibrated glucometer on arrival. In those known to be diabetic or those who had capillary glucose >7.8 mmol/L, we collected a fasting and post breakfast venous blood samples. In the remaining parents we performed 75g OGTT with a venous blood sample 2 hour after glucose load. OGTT results were interpreted using American Diabetes Association (ADA) 2014 criteria ^[19]^.

In children younger than 10 years of age, only a capillary blood glucose (fasting or non-fasting) was measured on a calibrated glucometer.

Venous plasma glucose, total and HDL cholesterol and triglyceride concentrations were measured using standard enzymatic methods on an autoanalyzer (Hitachi 902, Roche Diagnostics GmbH, Germany). LDL cholesterol was calculated using the Freidewald-Fredrickson formula. Measurements were subject to external quality control (EQAS) and all measurements had a co-efficient of variation (CV) <5%. Plasma insulin was measured using ELISA (Mercodia AB, SE-754 50 Uppsala, Sweden) with an intra- and inter-assay CV <7% (external quality control through UK-NEQAS). HbA1c was measured by HPLC (Bio-Rad D-10, Hercules, California).

We compared the performance of the glucometer (Alere Inc., Waltham, MA, United States) against the laboratory plasma glucose measurements on the fasting samples in the parents (n=162). The correlation between the two measurements (r= 0.930, p=0.000) was good across a wide range of concentrations (Supplementary figure 5a). The Bland Altman plot (Supplementary figure 5b) showed a negative bias of 0.19 mmol/L (95% CI −1.54-1.15), venous plasma levels being lower. This bias is lower than the admissible difference of 0.83 mmol/L (ISO Recommendation, ^[20]^).

In children >10 years, we used HOMA models to calculate insulin sensitivity (HOMA-S) and beta cell function (HOMA-β) using the iHOMA2 calculator (https://www.phc.ox.ac.uk/research/technology-outputs/ihoma2, last accessed August 2019) ^[21]^. We also calculated insulin sensitivity by the Matsuda Index ^[22]^, and insulin secretion by the insulinogenic index {modified to allow for non-linearity in the data, (ln{Insulin (30 min/fasting) /Glucose (30 min/fasting)} ^[23]^. We calculated disposition index as a measure of compensatory insulin secretion for prevalent insulin sensitivity, as [Insulinogenic index*Matsuda index] but used the log scale equation [Log (Insulinogenic index) + Log (Matsuda index)].

We measured anti Glutamic Acid Decarboxylase (GAD) and anti Zinc Transporter 8 (ZnT8) antibodies by ELISA (RSR Ltd. Llanishen, Cardiff, UK) in children who were diagnosed with prediabetes or diabetes, and in all children of type 1 diabetic mothers, to investigate islet autoimmunity.

### Maternal and offspring measurements during pregnancy and delivery

We obtained information from pregnancy records on the type of maternal diabetes (type 1, type 2 or GDM), maternal age, type of delivery and baby’s birthweight. Type 1 and type 2 diabetes were defined by ADA clinical criteria ^[19]^. GDM was diagnosed by WHO criteria till 2012 ^[24]^ and by IADPSG criteria thereafter ^[25]^. Small for gestational age (SGA) and large for gestational age (LGA) was defined using INTERGROWTH-21 study criteria ^[26]^. Control mothers provided this information by recall and by referring to their own medical records, if available.

### Statistical methods

We calculated the sample size considering body mass index (BMI) as the primary outcome. Estimated difference in BMI between ODMs and ONDMs was based on previously published literature from India ^[12,13]^. In the age group <10 years, sample size was calculated to detect 0.5 kg/m^2^ difference between the groups at 90% power and 5% significance level. This amounted to 108 participants in each group. For ≥10 years age group, sample size was calculated to detect 1.0 kg/m^2^ difference between the groups, at 90% power and 5% significance level, amounting to 77 participants in each group.

Data are presented as mean (SD) for normally distributed variables and as median (25^th^–75^th^ percentile) for skewed variables; skewed data was normalized before analysis. Given the wide age range, anthropometric and metabolic-endocrine measurements were converted to standard deviation (SD) scores by residual approach adjusting for age, gender and pubertal stage.

We were able to classify glucose tolerance by ADA criteria in parents and only in children >=10 years of age who underwent a full OGTT. For children below 10 years of age we had either a fasting or non-fasting capillary blood glucose for which there are no guidelines to classify glucose intolerance at this age. To be able to classify glucose intolerance for all children, we calculated separate SD scores for the three groups: 1. Those with fasting capillary glucose, 2. Those with non-fasting capillary glucose, and 3. Those with fasting venous plasma glucose. Those above median SD score were classified as glucose intolerant. Metabolic-endocrine characteristics of normoglycemic and hyperglycemic ODMs were compared by ANOVA, adjusting for their age, gender and BMI.

Independent associations between overweight-obesity and glucose intolerance in the offspring and corresponding parental characteristics were tested by multiple logistic regression adjusting for SES (combining diabetic and non-diabetic pregnancies). For this purpose parental exposures were classified as neither parent abnormal, one parent abnormal or both parents abnormal. Overweight-obesity for both parents was measured at follow up. For glucose intolerance, maternal status during pregnancy and paternal status at follow up were considered (Table 2). Prevalence of overweight-obesity and glucose intolerance in the children is shown in relation to corresponding parental phenotypes in Figures 2 and 3 (unadjusted).We used SPSS version 21.0 (IBM corporation, Armonk, NY) for analysis.

The study protocol was approved by the Institutional Ethics Committee of the KEM Hospital Research Centre (EC 1333/2014) and is registered with ClinicalTrials.gov (NCT03388723). Children over 18 years and parents signed an informed consent. For children below 18 years, we obtained parental consent, children between 12 and 18 years also signed an informed assent.

## Results

### Subjects

We reviewed records of 861 mothers treated for diabetes during pregnancy (71 type 1 diabetes, 81 type 2 diabetes and 709 GDM). We could contact 346 mothers, of whom 176 (20.4% of total) agreed to participate. Compared to 685 who could not be studied, these 176 women had similar age (29.4 vs. 29.5 years, p=0.827), BMI (24.1 vs. 25.2 Kg/m^2^, p=0.131), height (154.3 vs. 154.9 cm, p=0.433), glucose concentrations at diagnosis of GDM (FPG, 5.9 vs. 6.0 mmol/L, p=0.649, 2 hour glucose, 10.4 vs. 10.0, p=0.490) and birth weight of babies (2.82 vs. 2.89 Kg, p=0.191).

We studied a total of 200 ODMs (176 born in the index pregnancy and 24 younger siblings born in a subsequent diabetic pregnancy). Of 176 index ODMs, 133 were born in a GDM pregnancy, and 43 in a pre-gestational diabetic pregnancy (22 type 1 diabetes and 21 type 2 diabetes). Of 24 siblings, 20 were born to GDM, 3 to type 1 diabetic and 1 to type 2 diabetic mothers. We studied 176 mothers and 150 fathers of ODMs. In addition we studied 177 ONDMs and their parents (177 mothers, 163 fathers) as controls (Supplementary figure 1).

**Figure 1:**
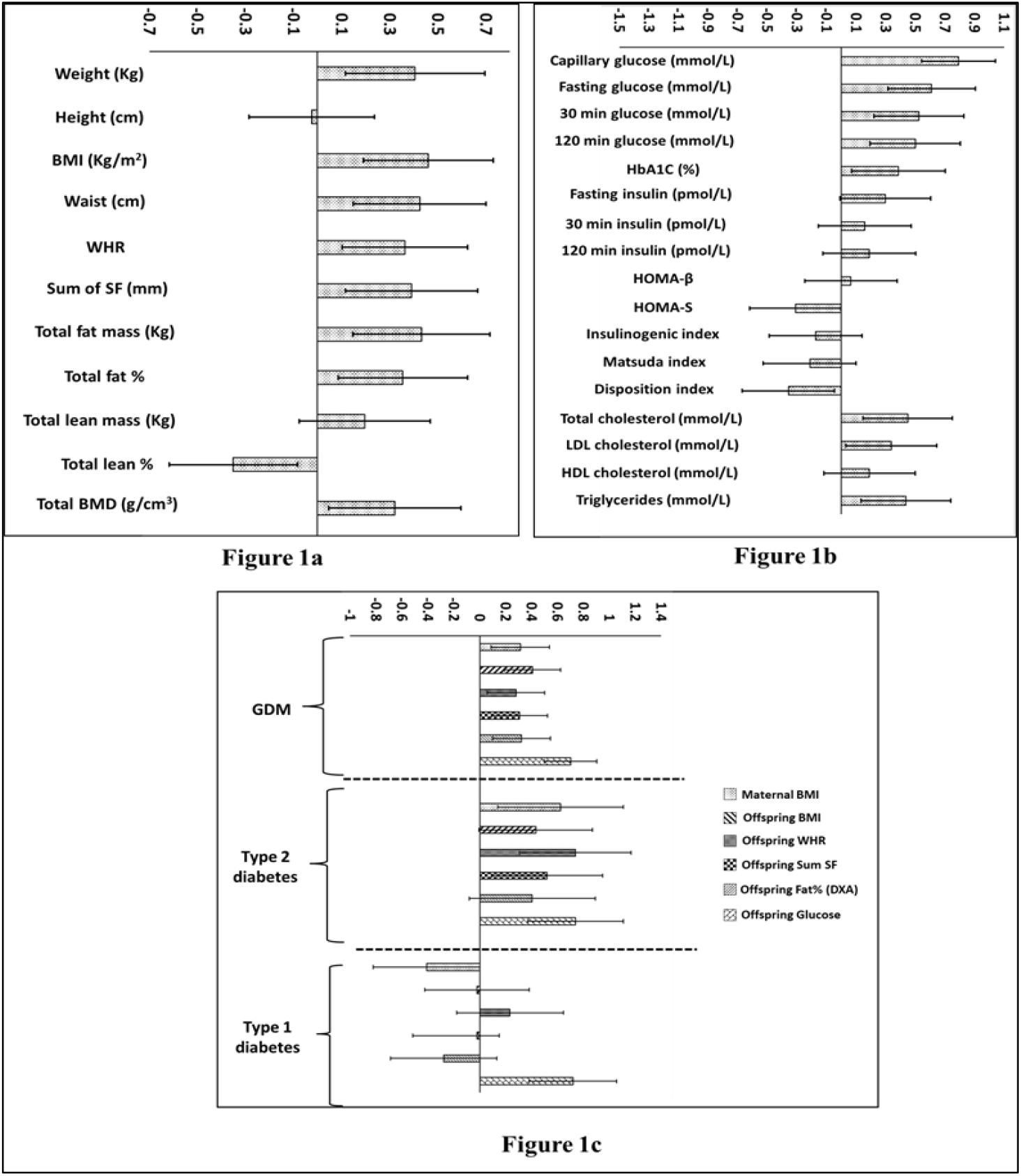
Differences between ODM and ONDM. Mean SD differences with 95% CI between ODM and ONDM (reference group) adjusted for age, gender and pubertal stage. Maternal BMI is in reference to control mothers. a): Anthropometric and DXA measurements b): Metabolic measurements c): Selected maternal and offspring characteristics by type of maternal diabetes (GDM, Type 2 diabetes, Type 1 diabetes) ODM= offspring of diabetic mothers, ONDM = offspring of non-diabetic mothers, BMI = Body mass index, WHR = Waist hip ratio, SF: Skin fold, HOMA-β: Homeostatic Model Assessment-Beta Cell function, HOMA-S: Homeostatic Model Assessment-Sensitivity.

### Comparison of ODM and ONDM

#### Pregnancy outcomes

ODMs had higher rates of preterm (34.4% vs. 19.9%, p=0.002) and caesarean deliveries (70.4% vs. 46.9%, p<0.001) compared to ONDMs. Birth weights of ODM and ONDM were comparable (mean 2.82 ± 0.64 kg and 2.85 ± 0.56 kg respectively, p=0.620). Prevalence of SGA was significantly lower (22.7% vs. 33.7, p=0.021) in ODM compared to ONDM but prevalence of LGA was similar (14.1% vs. 8.7%, p=0.101).

#### Follow up

At the time of study, ODM and ONDM had comparable distribution of age, sex ratio, pubertal stage and SES (supplementary table 1). Twenty ODM and 20 ONDM females had achieved menarche (at mean age of 12.2 ± 1.1 years and 12.2 ± 1.2 years respectively, p=0.947).

**Table 1:**
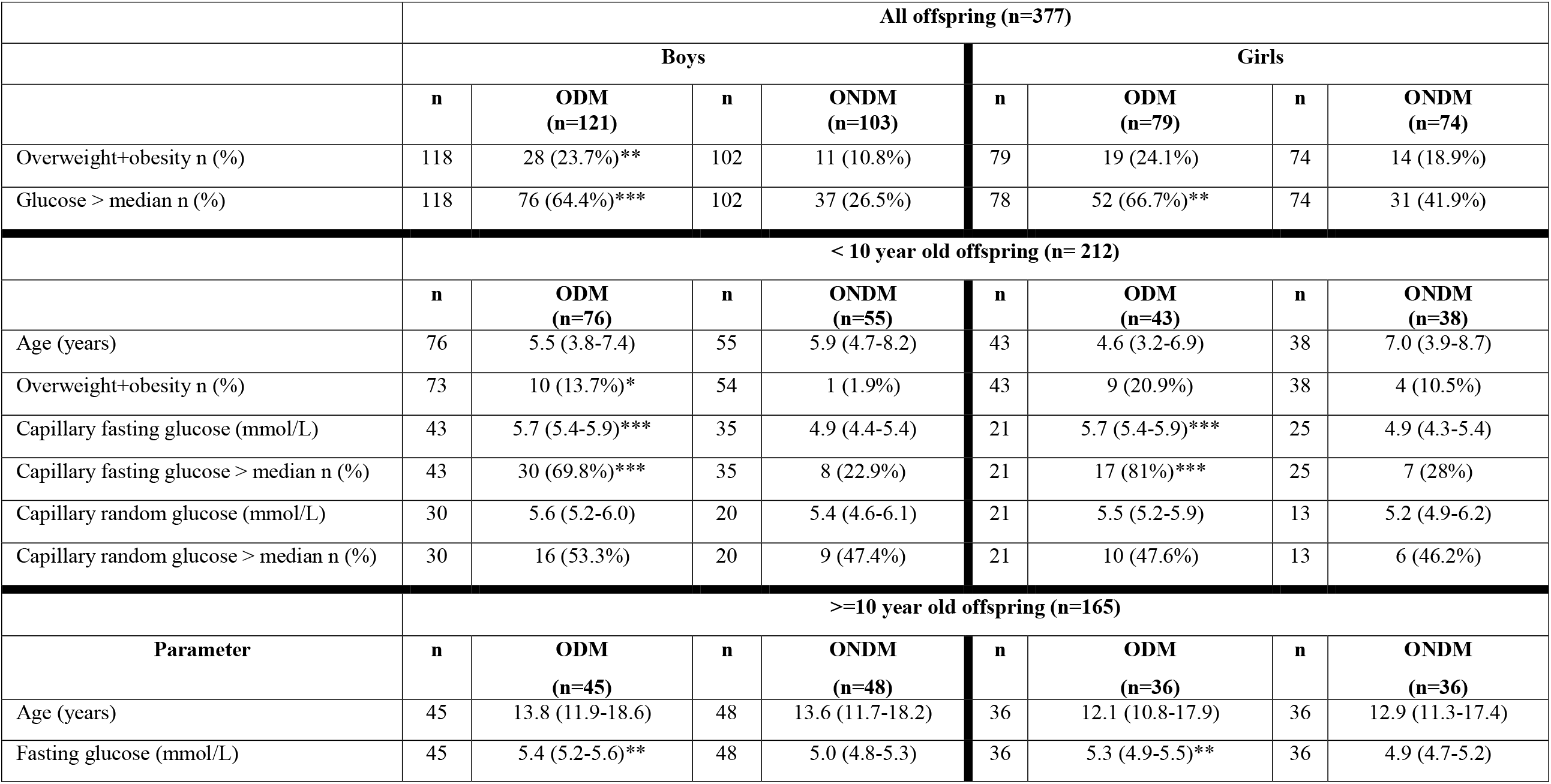
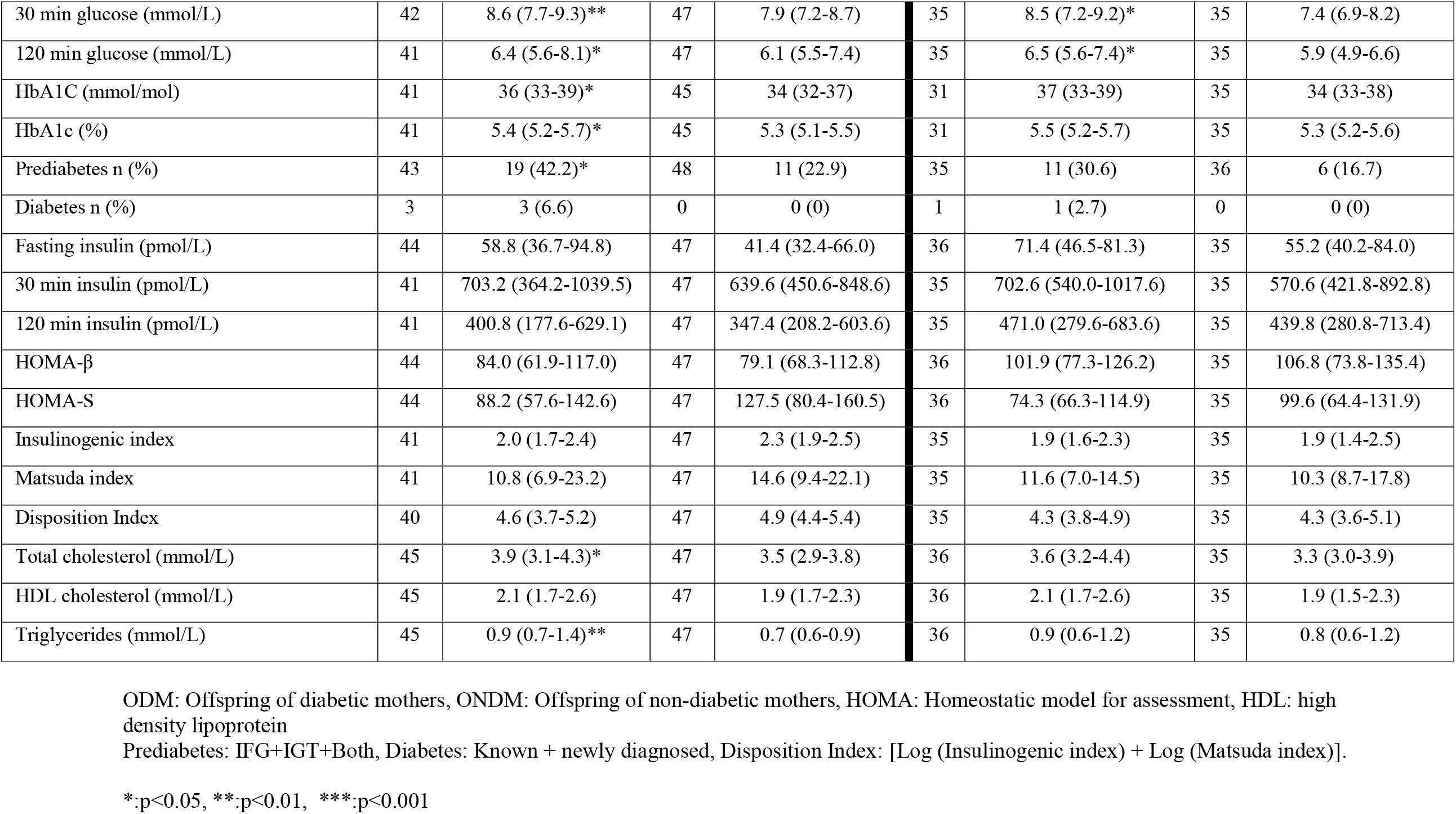
Characteristics of Offspring of Diabetic mothers (ODM) and Offspring of Non- Diabetic mothers (ONDMs)

##### Body size and composition

As a group, ODM had higher obesity related measurements (weight, BMI, WHR), and higher adiposity (body fat percent) compared to ONDM (Figure 1a). Prevalence of overweight + obesity (24% vs. 15%, p=0.014) was also higher in ODMs. When analysed separately by sex, anthropometric differences were significant only in males. (Supplementary Figure 2)

##### Metabolic parameters

###### Glycemia

Three ODM (>=10 years of age) were already receiving treatment for diabetes (diagnosed between 10-25 years of age). All were being treated with oral anti-diabetic agents and one also received insulin.

On OGTT, ODMs (>= 10 years) had higher prevalence of glucose intolerance (1 DM, 14 IFG, 12 IGT and 4 both IFG and IGT) compared to ONDM, (0 DM, 7 IFG, 9 IGT and 1 both IFG and IGT). All four diabetic ODMs were clinically classified as type 2 diabetes, and were born to GDM mothers. ODM also had higher HbA1c concentrations (Figure 1b). The glucose intolerant ODMs were more overweight+obese compared to normoglycemic ODMs (Supplementary Table 4). They also had higher triglyceride concentrations, and lower insulin sensitivity, lower disposition index and lower HDL concentrations adjusting for BMI.

Younger ODMs (<10 years) also had higher capillary blood glucose concentrations compared to ONDM (Table 1). This was driven by the group on whom fasting glucose was measured (5.7 vs 4.9 mmol/L, p=0.000, n=124), while the non-fasting glucose concentrations were similar in the two groups (5.6 vs 5.3 mmol/L, p=0.432, n=84).

Using the criterion of glucose concentration > median, ODMs had higher prevalence of glucose intolerance compared in ONDMs (65.2% vs. 32.8%), both in younger and older, and in boys and girls. (Table 1).

###### Glucose-insulin indices

HOMA-S and Disposition index were lower in ODMs compared to ONDMs. Other indices of insulin secretion and action (HOMA-β, Insulinogenic index and Matsuda index) were similar in the two groups (Figure 1b).

###### Lipids

ODM had higher total and LDL cholesterol and triglyceride concentrations compared to ONDM (Figure 1b).

###### Sex differences

As mentioned, anthropometric differences in ODMs and ONDMs were more prominent in males. ODM males also had a wider spectrum of metabolic abnormalities (higher concentrations of glucose, fasting insulin, HbA1c and triglycerides) while ODM females only had higher fasting and post challenge glucose concentrations compared to ONDMs. (Supplemental Figure 3).

###### Type of maternal diabetes, offspring body size and metabolism (Figure 1c)

Children of type 1 diabetic mothers had similar BMI, WHR and body fat percent (DXA) compared to ONDM, but had higher glycemia. On the other hand, children of GDM and type 2 diabetic mothers had higher BMI, WHR and glycemia. Children of GDM and type 2 diabetic mothers had higher concordance of overweight-obesity and glucose intolerance compared to children of type 1 diabetic mothers (∼20% versus 8.0% respectively).

###### Islet autoimmunity

None of the diabetic and pre-diabetic ODMs, not any of the children of type 1 diabetic mothers, were positive for circulating antiGAD or anti ZnT8 antibodies.

### Parental characteristics

#### Mothers

Mothers with type 2 diabetes and GDM (but not type 1 diabetes) were older and heavier than mothers of ONDM (Supplementary Table 2).

Eighty-one (61%) GDM mothers had diabetes at follow up (61 previously diagnosed, 20 newly diagnosed), and 32 (24%) had prediabetes. In comparison, 14 (8%) mothers of ONDM had diabetes and 46 (27%) had pre-diabetes. GDM mothers also had higher prevalence of overweight-obesity, central obesity and metabolic syndrome (74 vs. 33%) (Supplementary Table 3).

#### Father’s characteristics

Fathers of ODM had higher prevalence of prediabetes (49 vs. 33%, p=0.004) but similar prevalence of diabetes (17 vs 23%, p>0.05) compared to fathers of ONDM. Overweight-obesity and central obesity were similar in the two groups (Supplementary Table 2).

### Association of child’s overweight-obesity and glucose intolerance with parental characteristics

#### Overweight-obesity

There was an additive influence of maternal and paternal overweight-obesity on the phenotype of the child. The prevalence rose progressively with the number of parents being overweight-obese (none < one < both parents). This was true both for ODM and ONDM, though prevalence was higher in the ODM (Figure 2).

**Fig 2:**
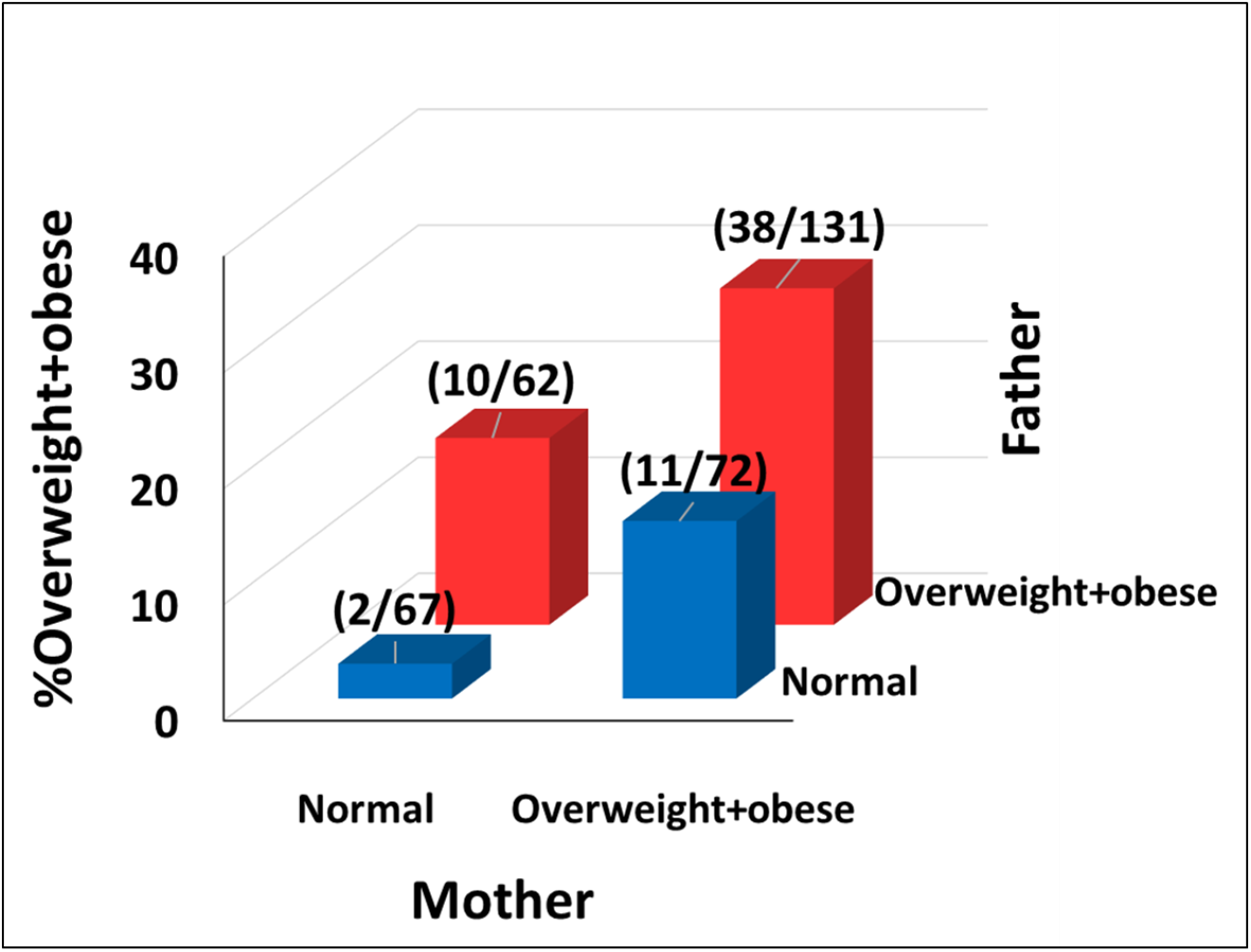
Overweight + obesity in offspring according to parental size. shows association between parental and offspring overweight-obesity, data from diabetic and non-diabetic pregnancies is combined. There is a progressive increase in the offspring overweight-obesity when none, one or both parents are overweight-obese. The figure highlights biparental transmission of overweight-obesity. Offspring: overweight-obesity was classified using IOTF (2-18 years) and WHO criteria (>18 years and parents). Overweight-obesity in parents ((WHO criteria, BMI >=25 Kg/m^2^) was measured at follow up. ODM: Offspring of diabetic mothers, ONDM: Offspring of non-diabetic mothers, IOTF: International Obesity Task Force, WHO: World Health Organization

#### Glucose intolerance

For glucose intolerance in the child, the picture was a bit more complex. By comparing the prevalence in the ODM and ONDM in relation to parental glucose intolerance we were able to compare the effect of intrauterine hyperglycemia against those of genetics and family environment. The lowest prevalence of glucose intolerance was in the ONDM when both parents did not have glucose intolerance. Maternal postnatal glucose intolerance made little difference, paternal glucose intolerance increased the risk. The highest prevalence was in the ODM children who were exposed to intrauterine hyperglycemia irrespective of paternal glucose intolerance (Figure 3).

**Fig 3:**
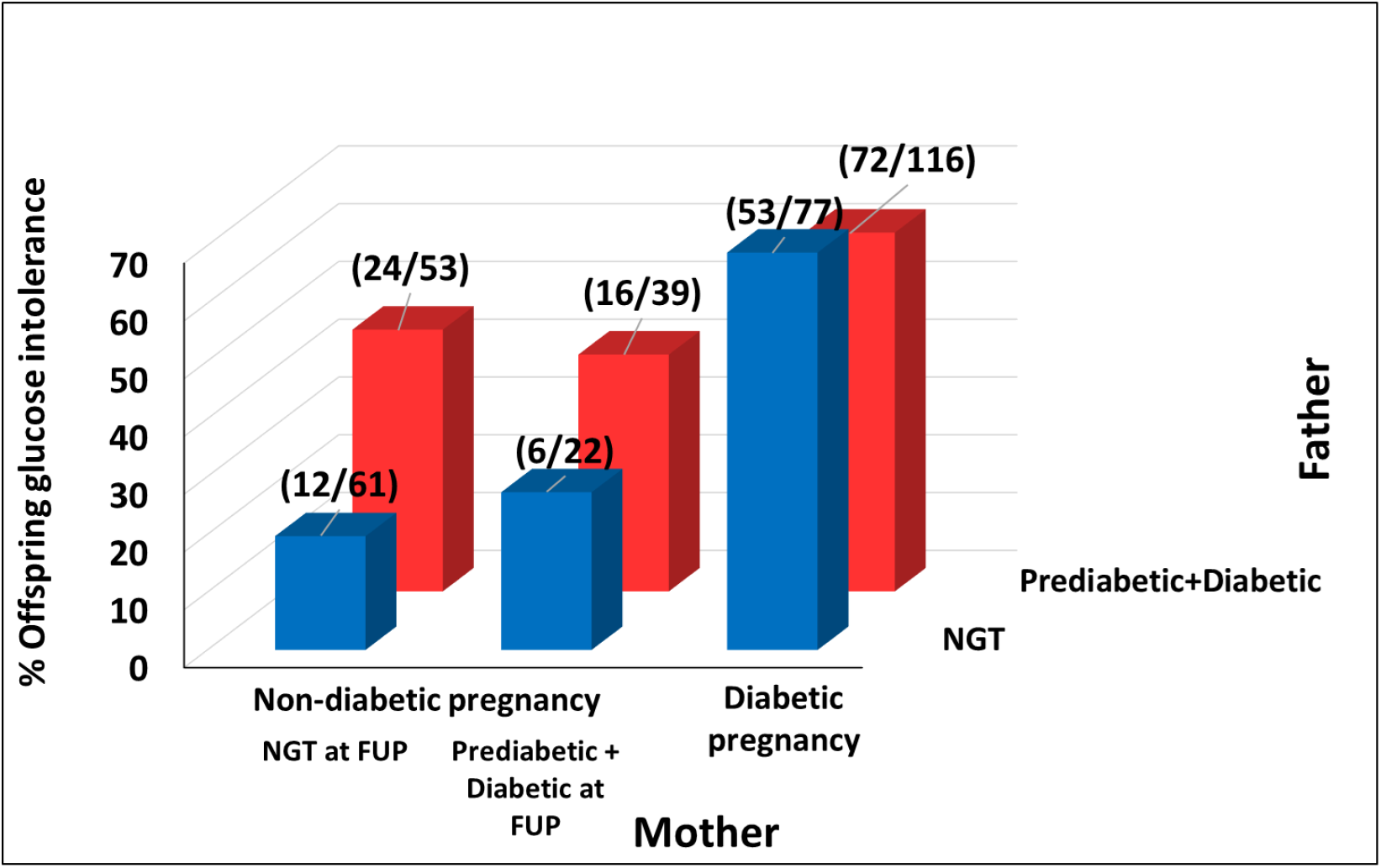
Glucose intolerance in offspring according to parental glycemia. shows association between parental-child glucose intolerance. When mother is not diabetic during pregnancy or subsequently, the children have lower risk of glucose intolerance. Children of mothers who developed glucose intolerance subsequently (i.e were ‘pre-diabetic’ in pregnancy) also have similar low risk. Paternal diabetes increases child’s risk of glucose intolerance in ONDM irrespective of mother becoming glucose intolerant after delivery. When mother has pregnancy diabetes the children have high risk of glucose intolerance which is not increased by glucose intolerant fathers. Thus, maternal diabetes in pregnancy is the strongest risk factor for glucose intolerance in the child, highlighting a role for intrauterine environment. Glucose intolerance was defined in offspring as glucose concentration above median for the age group. In parents it was based on ADA 2014 criteria (pre-diabetes+diabetes). ODM: Offspring of mothers who had diabetes in pregnancy, ONDM: Offspring of mothers who did not have diabetes in pregnancy, ADA: American Diabetes Association

Multivariate analysis showed that the influence of parental overweight-obesity on child’s overweight obesity was independent of parental glucose intolerance. Similarly, the influence of parental glucose intolerance on child’s glucose intolerance was independent of parental overweight-obesity. We interpret this as a ‘mirror imaging’ of overweight-obesity and glucose-intolerance between parents and children. Child’s birth weight was positively associated only with risk of later overweight-obesity and not with glucose intolerance. SES was not associated with either outcome (Table 2).

**Table 2:**
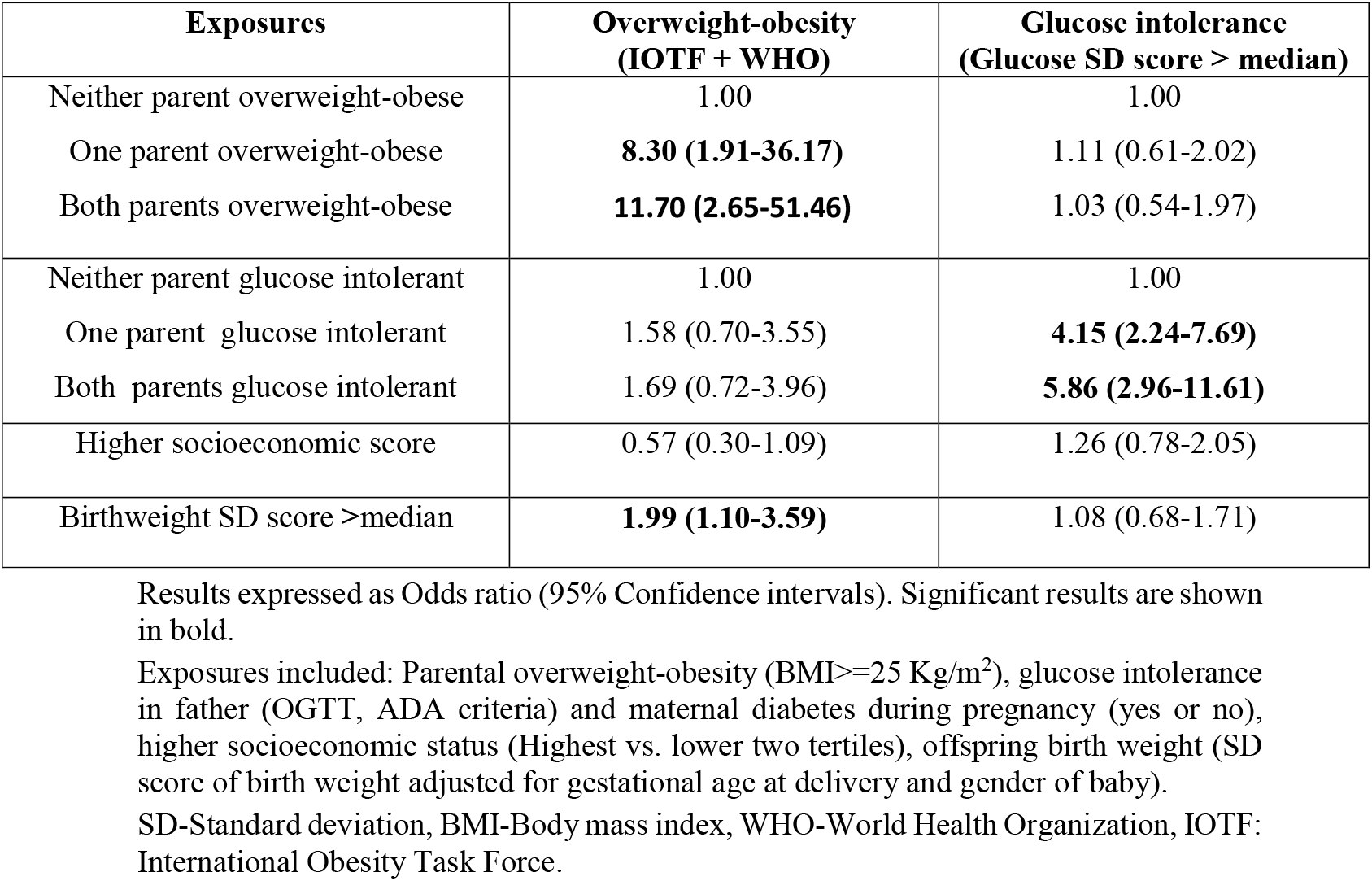
Multivariate Logistic Regression for factors associated with overweight-obesity and glucose intolerance in offspring (ODM + ONDM, n=373)

## Discussion

Our results confirm that the children born in a diabetic pregnancy are more likely to be overweight-obese and glucose intolerant than those born in a non-diabetic pregnancy. These phenotypes were apparent from early childhood. Overweight-obesity and glucose intolerance were not necessarily concordant, suggesting an independence of the two phenotypes. Children of thin type 1 diabetic mothers, despite their exposure to much higher glucose levels *in utero*, were thinner than the children of heavier mothers with GDM and type 2 diabetes. Maternal diabetes during pregnancy had an overriding influence on risk of glucose intolerance of the child, paternal glycemia added little. For overweight-obesity, paternal size added to the maternal effect. Birthweight was associated only with risk of later overweight-obesity but not to glucose intolerance.

Our results indicate a bi-parental influence on post-natal overweight-obesity in ODM that is uninfluenced by parental glycemia, including intrauterine hyperglycemia. The effect appears to be specific for adiposity, given that the height and lean mass of ODMs and ONDMs were similar. The only comparable Indian follow up study (the Parthenon Cohort) made serial anthropometric measurements in ODM and ONDM and found that the ODMs weighed more at birth, ‘caught-down’ during infancy and showed a rebound in early childhood, especially in females ^[12, 13]^. We observed the effects predominantly in males. European studies have reported increased adiposity in both sexes or restricted to males ^[27]^. It would be interesting to explore the sex differences further.

The ODMs had higher glucose and HbA1c concentrations than the ONDMs. Early-onset type 2 diabetes (<20 years) was seen only in the ODM. Parental glycemia but not overweight-obesity was associated with child’s glycemia. Of interest, and somewhat contrary to popular belief, birth weight was unrelated to glucose intolerance. These findings suggest that glucose intolerance and overweight are ‘programmed’ independently (genetically and/or epigenetically), and that the reported high concordance rate for these two conditions in ODMs may simply reflect the frequent coexistence of these two conditions in the parents. Only 8% of children born to type 1 diabetic mothers had both glucose intolerance and overweight-obesity, compared to ∼20% born to GDM or type 2 diabetic mothers.

Several studies have reported an increased prevalence of glucose intolerance in the offspring born in diabetic pregnancies ^[3, 12, 13]^. There is ongoing debate about the relative contribution of genetics, intra-uterine environment and lifestyle to this risk. The best evidence for a predominant role of intra-uterine epigenetic programming comes from studies in Pima Indians in whom children born in diabetic pregnancies had a higher risk of diabetes (and obesity) compared to those born in non-diabetic pregnancies or siblings born before the mother became diabetic (therefore, genetically predisposed but not exposed to intrauterine hyperglycemia), and to children born to diabetic fathers (therefore, genetically predisposed or sharing family environment) ^[28, 29]^. Pregnancy diabetes is estimated to contribute up to 40% of diabetes in the young Pima Indians ^[30]^. Our results replicate these findings. Children born in a diabetic pregnancy had the highest prevalence of glucose intolerance. The young diabetic children were all born in a diabetic pregnancy. Paternal glucose intolerance did not add to this effect. Norbert Freinkel called intrauterine non-genetic transmission ‘fuel mediated teratogenesis’ referring to macrosomia and later obesity and diabetes ^[31]^.

Glucose intolerance in our ODMs can be attributed to a contribution of lower insulin sensitivity and lower insulin secretion in relation to prevailing insulin insensitivity (i.e. a lower disposition index) rather than to an absolute reduction in insulin secretion. Relatively higher reduction in fasting insulin sensitivity (HOMA-S) compared to post glucose challenge measurement (Matsuda index) could be interpreted as higher contribution from adipose and hepatic than muscle insulin insensitivity. Higher triglycerides in ODMs also support such a pathophysiology.

Follow up studies in children from the Hyperglycemia and Adverse Pregnancy Outcomes (HAPO) study have reported independent associations of maternal BMI and pregnancy glycemia (across the range) with offspring obesity-adiposity and glycemia, respectively ^[6–8]^. Maternal overweight-obesity were more strongly associated with the child’s adiposity than maternal glycemia and vice versa ^[9]^. HAPO also showed a mediating role for birth size in the association between maternal BMI and a child’s BMI ^[10]^. However they lack paternal data (other than history of diabetes) nor did they analyze discordant maternal exposures (thin diabetic mothers or obese-low glucose mothers). Our study is much smaller, but enabled us to explore these interesting exposures especially due to inclusion of type 1 diabetic pregnancies.

### Strengths and limitations

Ours is the largest Indian follow up of children born in diabetic pregnancies. Relative thinness of diabetic mothers compared to western studies and inclusion of type 1 diabetic pregnancies provided an opportunity to disentangle the effects of maternal obesity and glucose intolerance on offspring phenotypes. Studying fathers helped to define their role in a condition usually ascribed only to maternal diabetes. The wide age range in ODMs revealed that abnormalities in body size and metabolism are manifest from early childhood. Only a fifth of eligible women could be followed up. However, their pregnancy characteristics (mothers and babies) were similar to those who could not be studied, thus considered representative. As a hospital-based study, there may have been some selection bias. However, the KEM Hospital attracts people from all socio-economic classes, and all those with diagnosed diabetes in pregnancy are usually managed in hospital settings. Changing criteria for diagnosis of GDM during the study period may have introduced some heterogeneity. The control group was mostly classified based only on recall but the low prevalence of glucose intolerance at follow up in mothers argues against substantial misclassification. We could only perform capillary glucose measurements in children less than 10 years of age. This was a pragmatic and ethical decision owing to parents’ discomfort in subjecting younger children to venipuncture and multiple sampling in a standard OGTT. However, there was a good correlation between glucometer and laboratory glucose readings and the bias was within allowable analytical error.

In summary, we confirm the increased long-term risk of overweight+obesity and glucose intolerance in Indian ODMs. There was an independent prediction of these two outcomes through corresponding phenotypes of the parents. Interestingly, there was a biparental contribution to the risk of overweight+obesity but maternal pregnancy diabetes was the overriding predictor of offspring glucose intolerance. Future genetic and epigenetic studies will help better understand the basis of fetal programming in diabetic pregnancies.

## Data Availability

Data is available with Prof. C.S.Yajnik for sharing to confirm our findings and for additional analysis by applying to the corresponding author with a 200 word plan of analysis. Data sharing is subject to KEMHRC, Ethics Committee approval and Govt. of India Health Ministry Advisory Committee permission.

## Acknowledgements

This investigation was part of the InDiaGDM grant of the Department of Biotechnology, New Delhi, India (BT/IN/Denmark/02/CSY/2014).

We are grateful to Prof. Edwin Gale for informative discussions. We also acknowledge Neelam Memane, Deepa Raut, Dr. SDK, Dr. Smita Dhadge, Dr. Meenakumari, Aboli Bhalerao, Swati Alekar, Alma Baptist, Preeti Kalel, Ajay, Abhay, Dr. Rohan Shah, Rupali Joshi and Dr. Giriraj Chandak from CCMB (co-investigator in InDiaGDM study) for their valuable contributions to the study.

## Author contributions

SW, KK, MD, CSY were involved in study conceptualization, subject recruitment and actual conduct of the study. SW, SR, RL, PCY were involved in study administration. SW, MD were involved in data curation and formal analysis. DB, RK and Sayali Wadke were involved in laboratory analysis. SW, SP and CSY were involved in visualization, writing, reviewing and the manuscript.

## Data sharing statement

Data is available with Prof. C.S.Yajnik by applying with a 200 word plan of analysis. Data sharing is subject to KEMHRC Ethics Committee approval and Govt. of India’s Health Ministry Advisory Committee permission.

