## Supplemental tables and figures for "Overweight-Obesity And Glucose Intolerance In Offspring Of Indian Diabetic Mothers"

**Supplemental material: Index**


| Sr No. | Item | Pg. No. |
|---|---|---|
| 1 | Supplemental Table S1: Anthropometry and body composition of offspring (ODM andONDM) at the time of study | 2-3 |
| 2 | Supplemental Table S2: Measurements of mothers (pregnancy and follow up) and fathers(follow up) | 4-5 |
| 3 | Supplemental Table S3: Maternal measurements in GDM, T2D and T1D mothers at followup | 6 |
| 4 | Supplemental Table S4: Biochemical characteristics of Hyperglycemic vs.Normoglycemic ODMs (>=10 years) | 7 |
| 5 | Supplementary figure 1: Flow diagram of the study | 8 |
| 6 | Supplementary figure 2: Gender wise differences for anthropometric and DXA measurements between ODM and ONDM | 9 |
| 7 | Supplementary figure 3: Gender wise differences for metabolic measurements between ODM and ONDM | 10 |
| 8 | Supplementary figure 4: Overweight + obesity in offspring according to parental size in ODM and ONDM | 11 |
| 9 | Supplementary figure 5: Agreement between venous plasma glucose and capillary glucose in parents | 12 |

**Supplementary Table 1: Anthropometry and body composition of offspring (ODM and ONDM) at the time of study**

| Parameter | Boys | | Girls | |
|---|---|---|---|---|
| | **ODM (n=121)** | **ONDM (n=103)** | **ODM (n=79)** | **ONDM (n=74)** |
| Age (years) | 8.0 (4.6-12.3) | 9.1 (5.6-13.0) | 8.4 (4.1-12.0) | 9.7 (6.8-12.9) |
| SLI | 35.0 (40.0-43.0) | 34.8 (38.0-42.0) | 35.0 (38.0-42.8) | 34.0 (39.0-43.0) |
| Height (cm) | 126.8 (105.5-154.9) | 130.7 (112.7-154.1) | 128.0 (104.9-152.1) | 136.5 (113.9-152.5) |
| Weight (Kg) | 25.0 (15.8-47.6) | 26.8 (17.7-44.9) | 27.8 (15.7-43.9) | 31.1 (17.6-45.8) |
| BMI (Kg/m$^2$) | 16.0 (14.2-20.9) | 15.7 (14.1-18.5) | 16.8 (14.4-20.1) | 16.6 (14.0-20.1) |
| Waist (cm) | 57.8 (49.6-78.3) | 59.5 (51.0-72.4) | 61.2 (51.0-73.4) | 60.0 (50.3-74.8) |
| Hip (cm) | 69.0 (54.3-88.9) | 68.4 (57.2-84.0) | 70.0 (56.0-85.0) | 73.9 (58.9-89.8) |
| WHR | 0.90 (0.86-0.94) | 0.86 (0.83-0.91) | 0.87 (0.81-0.92) | 0.84 (0.80-0.89) |
| MUAC (cm) | 19.6 (16.2-25.4) | 18.8 (16.0-23.9) | 20.3 (16.1-24.2) | 20.7 (16.6-24.4) |
| Bicep (mm) | 6.4 (4.5-9.1) | 5.5 (3.9-7.7) | 6.7 (5.0-9.4) | 7.1 (5.1-10.4) |
| Tricep (mm) | 10.3 (7.4-15.6) | 9.2 (7.3-13.9) | 11.2 (8.6-16.9) | 12.2 (8.5-18.4) |
| Subscapular (mm) | 9.2 (5.8-19.9) | 7.9 (5.4-16.3) | 10.2 (7.0-22.9) | 13.1 (7.1-28.8) |
| Suprailiac (mm) | 11.9 (6.4-30.9) | 9.4 (6.2-24.9) | 19.0 (8.8-36.7) | 15.8 (8.2-30.5) |
| Sum of skinfolds (mm) | 36.9 (25.4-80.0) | 32.3 (23.5-65.6) | 44.6 (29.9-88.4) | 48.5 (30.1-88.1) |
| **Whole body composition (DXA)** | | | | |
| | **ODM (n=110)** | **ONDM (n=99)** | **ODM (n=71)** | **ONDM (n=69)** |
| Fat mass (Kg) | 5.8 (2.3-15.7) | 4.2 (2.3-10.9) | 8.5 (3.3-15.5) | 8.3 (3.1-15.6) |
| Total fat % | 20.5 (13.9-31.8) | 17.3 (11.6-28.1) | 27.0 (19.8-38.8) | 27.3 (17.7-38.7) |

| | | | | |
|---|---|---|---|---|
| Lean mass (Kg) | 19.8 (13.6-33.4) | 19.0 (14.1-31.3) | 19.2 (12.8-28.1) | 20.8 (15.5-27.1) |
| Total lean % | 76.1 (64.6-82.8) | 78.7 (68.0-84.7) | 69.4 (58.0-77.2) | 68.9 (57.9-78.9) |
| Total BMD (g/cm$^3$) | 0.840 (0.750-0.957) | 0.844 (0.767-0.943) | 0.837 (0.734-0.979) | 0.833 (0.759-0.959) |
| Spine BMD (g/cm$^3$) | 0.657 (0.560-0.796) | 0.668 (0.568-0.763) | 0.698 (0.573-0.869) | 0.690 (0.586-0.883) |

Values are in Median (25$^{th}$-75$^{th}$ centile), ODM: Offspring of diabetic mothers, ONDM: Offspring of non-diabetic mothers, SLI: Standard of living index, BMI: Body mass index, MUAC: Mid upper arm circumference, WHR: Waist to hip ratio, BMD: Bone mineral density
Given the wide range of age we have not calculated the significance between ODM and ONDM in this table. Figure 1a shows age, gender and pubertal status standardized scores of these characteristics for statistical comparison.

**Supplementary Table 2: Measurements of mothers (pregnancy and follow up) and fathers (follow up)**

| **Mothers** | | | |
|---|---|---|---|
| **Measurements** | **GDM (n=133)** | **Non-GDM (n=177)** | **p** |
| **Pregnancy** | | | |
| Pre-pregnancy weight (kg) | 57.0 (50.0-63.0) | 50.0 (45.0-57.0) | **<0.001** |
| Age at delivery (y) | 29.6 (25.8-32.7) | 25.7 (22.6-28.4) | **<0.001** |
| Multiparity n (%) | 89 (58.6) | 89 (50.3) | 0.082 |
| Gestation at delivery (weeks) | 38.0 (36.6-39.0) | 39.4 (37.4-40.1) | **<0.001** |
| Caesarean deliveries n (%) | 105 (55.9) | 83 (44.1) | **<0.001** |
| Preterm deliveries n (%) | 47 (31.1) | 35 (19.9) | **0.014** |
| Offspring Birth weight (gms) | 2850.0 (2521.1-3300.0) | 2830.0 (2500.0-3250.0) | 0.829 |
| **Follow up** | | | |
| **Anthropometry** | **GDM (n=133)** | **Non-GDM (n=177)** | **p** |
| Age (years) | 38.4 (34.5-44.2) | 35.1 (31.1-39.6) | **<0.001** |
| Years since index pregnancy | 9.7 (5.6) | 10.8 (5.6) | 0.228 |
| Height (cm) | 154.2 (149.8-156.2) | 155.0 (151.5-158.2) | 0.245 |
| Weight (kg) | 62.8 (56.7-71.6) | 60.7 (54.1-70.2) | 0.066 |
| BMI (Kg/m$^2$) | 27.1 (23.9-30.6) | 25.8 (22.7-28.7) | **0.002** |
| Overweight + obesity n (%) | 91 (68.9) | 101 (57.4) | **0.038** |
| Waist circumference (cm) | 91.0 (85.7-98.7) | 89.1 (82.3-97.3) | **0.003** |
| Hip (cm) | 103.0 (95.2-111.2) | 101.4 (95.0-107.9) | **0.003** |
| WHR | 0.89 (0.86-0.93) | 0.88 (0.83-0.91) | **0.003** |
| Central obesity n (%) (WHR >0.8) | 126 (94.7) | 149 (84.1) | **0.003** |
| **Glycemic status** | | | |
| Diabetes n (%) | | | |
| • Known diabetes | 61 (46%) | 7 (4%) | |
| • Diagnosed at follow up | 20 (15%) | 5 (3%) | |
| • Total diabetes | 81 (61%) | 12 (7%) | **<0.001** |
| Pre-diabetes n (%) | 32 (24%) | 46 (27%) | 0.699 |
| Normal Glucose Tolerance (NGT) n (%) | 20 (15%) | 117 (66%) | **<0.001** |

| Fathers | | | |
|---|---|---|---|
| **Measurements** | **n=150** | **n=163** | |
| Age (years) | 41.8 (37.5-45.9) | 39.7 (36.2-45.1) | **0.021** |
| Height (cm) | 168.2 (164.6-173.1) | 167.7(163.9-173.5) | 0.716 |
| Weight (kg) | 73.2 (65.9-84.7) | 72.0 (64.5-80.7) | 0.111 |
| BMI (Kg/m2) | 25.8 (23.5-28.7) | 25.3 (23.2-27.6) | 0.114 |
| Overweight + obesity n (%) | 88 (59.1) | 88 (54.3) | 0.404 |
| Waist (cm) | 96.9 (90.1-104.0) | 95.6 (89.3-101.5) | 0.158 |
| Hip (cm) | 100.2 (34.5-104.5) | 98.3 (95.2-104.0) | 0.253 |
| WHR | 0.97 (0.92-1.00) | 0.96 (0.92-0.99) | 0.678 |
| Central obesity n (%) (WHR >0.9) | 130 (86.6) | 148 (90.7) | 0.246 |
| **Glycemic status** | | | |
| Diabetes n (%)<br>• Known diabetes<br>• Diagnosed at follow up<br>• Total diabetes | <br>10 (6.6)<br>16 (10.6)<br>26 (17.3) | <br>15 (9.2)<br>23 (14.1)<br>38 (23.3) | <br><br><br>0.190 |
| Pre-diabetes n (%) | 73 (48.6) | 53 (32.5) | **0.003** |
| Normal Glucose Tolerance (NGT) n (%) | 49 (32.6) | 72 (44.2) | **0.036** |

Values are in median ($25^{th}$-$75^{th}$ percentile) or n (%), BMI: Body mass index, WHR: Waist to hip ratio, NGT: Normal glucose tolerance, GDM: Gestational Diabetes Mellitus

**Supplementary Table 3: Maternal measurements in GDM, T2D and T1D mothers at follow up**

| **Measurements** | **GDM (n=133)** | **T2D (n=21)** | **T1D (n=22 )** | **Non-diabetic (n=177)** | **p1** | **p2** | **p3** |
|---|---|---|---|---|---|---|---|
| Age (years) | 38.4 (34.5-44.2) | 37.3 (34.5-38.9) | 32.8 (30.5-40.9) | 35.1 (31.1-39.6) | **<0.001** | 1.000 | 1.000 |
| Height (cm) | 154.2 (149.8-156.2) | 154.8 (150.4-159.2) | 157.5 (150.9-162.4) | 155.0 (151.5-158.2) | 0.245 | 1.000 | 1.000 |
| Weight (kg) | 62.8 (56.7-71.6) | 68.9 (62.7-84.4) | 55.1 (52.7-61.9) | 60.7 (54.1-70.2) | 0.066 | **0.017** | 1.000 |
| BMI (Kg/m2) | 27.1 (23.9-30.6) | 28.7 (27.2-32.7) | 22.9 (22.1-25.3) | 25.8 (22.7-28.7) | **0.002** | **0.013** | 0.427 |
| Waist (cm) | 91.0 (85.7-98.7) | 100.8 (94.5-107.1) | 83.7 (81.3-91.0) | 89.1 (82.3-97.3) | **0.003** | **0.001** | 0.837 |
| Hip (cm) | 103.0 (95.2-111.2) | 107.5 (101.3-118.0) | 98.0 (97.0-100.5) | 101.4 (95.0-107.9) | **0.003** | **0.001** | 1.000 |
| WHR | 0.89 (0.86-0.93) | 0.92 (0.89-0.97) | 0.85 (0.83-0.89) | 0.88 (0.83-0.91) | **0.003** | **<0.001** | 1.000 |
| Central obesity n (%) (WHR >0.8) | 126 (94.7) | 17 (80.9) | 18 (81.8) | 149 (84.1) | **0.003** | 0.703 | 0.776 |
| Overweight+obesity n (%) | 91 (68.9) | 17 (81.0) | 8 (36.4) | 101 (57.4) | **0.038** | **0.037** | 0.062 |
| Systolic BP (mmHg) | 109.0 (103.5-116.5) | 115.5 (104.0-120.0) | 107.5 (97.0-118.0) | 106.5 (100.5-114.0) | 0.245 | 0.416 | 1.000 |
| Diastolic BP (mmHg) | 71.5 (67.0-76.0) | 74.0 (67.5-82.5) | 69.0 (65.5-70.0) | 69.5 (64.5-74.5) | 0.186 | 0.409 | 1.000 |
| Pulse (/min) | 78.5 (73.0-86.0) | 85.0 (73.5-95.5) | 82.5 (73.0-93.0) | 75.5 (68.0-81.5) | **0.001** | **<0.001** | **0.004** |

Median ($25^{th}$-$75^{th}$ centile) for continuous variables n (%) for categorical variables, BMI: Body mass index,
WHR: Waist to hip ratio, GDM: Gestational Diabetes Mellitus
p1: GDM Vs Non-diabetic, p2: T2D Vs Non-diabetic, p3: T1D Vs Non-diabetic

**Supplementary Table 4: Metabolic-endocrine characteristics of Hyperglycemic vs. Normoglycemic ODMs (>=10 years) based on OGTT**

| Measurements | Hyperglycemic (n=31) | Normoglycemic (n=47) | p1 | p2 |
|---|---|---|---|---|
| Boys n (%) | 20 (64.5) | 23 (48.9) | **--** | **--** |
| Age (years) | 14.2 (12.0-17.9) | 12.2 (11.1-16.8) | 0.526 | -- |
| BMI (Kg/m$^2$) | 21.8 (18.8-26.6) | 19.4 (16.8-22.9) | **0.023** | -- |
| Total body fat (%) | 35.2 (26.4-42.9) | 29.8 (23.3-37.5) | **0.019** | -- |
| Fasting glucose (mmol/L) | 5.61 (5.40-5.76 ) | 5.22 (5.05-5.33) | **0.000** | **0.000** |
| 30 min glucose (mmol/L) | 9.05 (8.33-9.72) | 8.11 (7.22-8.77) | **0.000** | **0.000** |
| 120 min glucose (mmol/L) | 8.00 (6.72-9.22) | 6.22 (5.47-6.63) | **0.000** | **0.000** |
| HbA1c (%) | 5.6 (5.3-5.8) | 5.3 (5.1-5.5) | **0.004** | **0.020** |
| Fasting insulin (pmol/L) | 79.2 (48.0-116.4) | 54.3 (36.4-81.9) | **0.006** | **0.000** |
| 30 min insulin (pmol/L) | 825.6 (510.6-1207.8) | 593.4 (400.5-959.7) | **0.044** | 0.121 |
| 120 min insulin (pmol/L) | 572.4 (308.4-1500.0) | 291.6 (174.9-494.4) | **0.000** | **0.003** |
| HOMA-β | 102.5 (69.7-125.9) | 91.1 (71.7-110.5) | 0.158 | **0.000** |
| HOMA-S | 67.2 (45.8-109.8) | 97.7 (65.8-144.4) | 0.214 | 0.258 |
| Insulinogenic index | 1.94 (1.66-2.30) | 2.03 (1.69-2.37) | 0.778 | 0.992 |
| Matsuda index | 8.1 (3.7-14.2) | 13.8 (9.3-23.1) | **0.000** | **0.001** |
| Disposition Index | 4.05 (2.85-4.55) | 4.65 (4.18-5.18) | **0.002** | **0.010** |
| Total cholesterol (mmol/L) | 3.98 (3.08-4.48) | 3.60 (3.13-4.06) | 0.728 | 0.706 |
| LDL cholesterol (mmol/L) | 2.38 (1.68-2.72) | 1.97 (1.72-2.27) | 0.593 | 0.634 |
| HDL cholesterol (mmol/L) | 1.03 (0.93-1.11) | 1.13 (0.98-1.34) | **0.020** | 0.067 |
| Triglycerides (mmol/L) | 1.11 (0.92-1.41) | 0.87 (0.57-1.07) | **0.005** | **0.015** |

Values in median (25$^{th}$-75$^{th}$ percentile), p1: adjusted for age and gender except for age, p2: adjusted for age, gender and child's BMI except for age

ODM: Offspring of diabetic mothers, ONDM: Offspring of non-diabetic mothers, OGTT: Oral glucose tolerance test, HOMA: Homeostatic model for assessment, LDL: low density lipoprotein, HDL: high density lipoprotein, Disposition Index: [(Insulinogenic index + log (Matsuda index)]

**Supplementary figure 1: Flow diagram of the study**

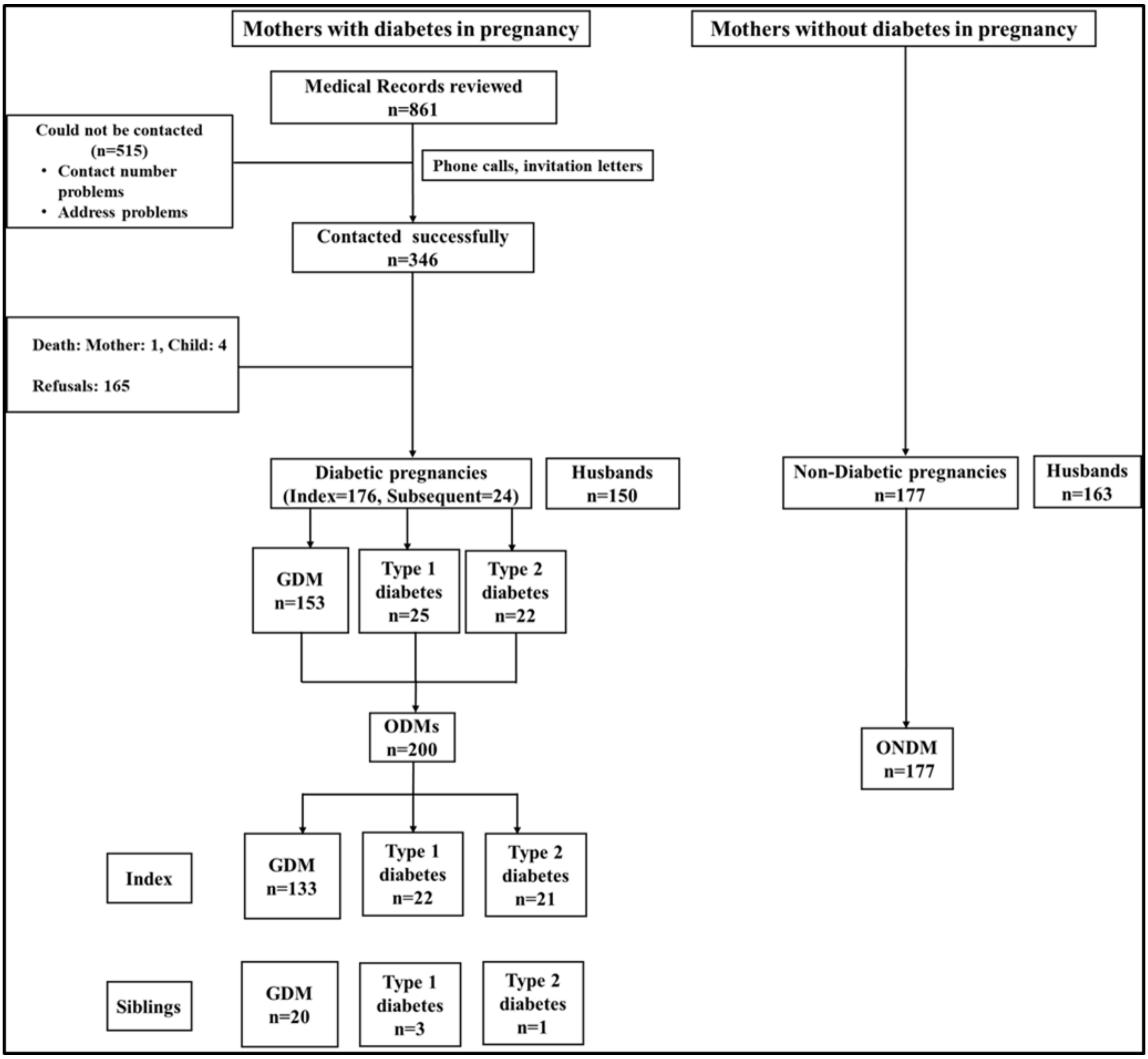


Supplementary Figure 1: Flow diagram of the study. Women attending pregnancy diabetes clinic were contacted to participate in the follow up of their children. Families who consented were studied. Control children whose mothers were not diabetic during pregnancy were studied along with their parents.
ODM: Offspring of diabetic mothers, ONDM: Offspring of non-diabetic mothers, GDM: Gestational Diabetes Mellitus

**Supplementary figure 2: Gender wise differences for anthropometric and DXA measurements between ODM and ONDM**

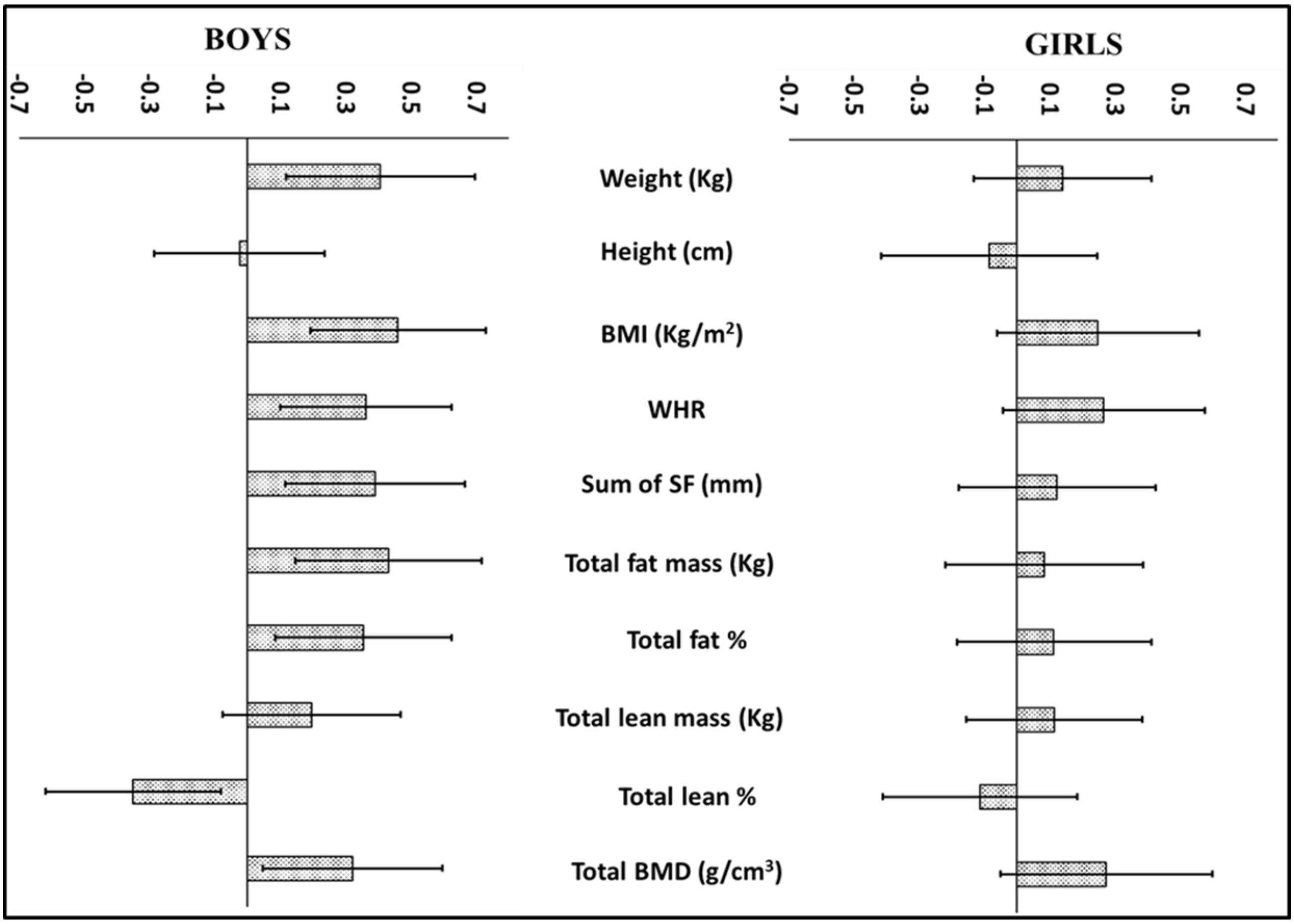


Supplementary Figure 2: Mean difference with 95% CI of SD scores for anthropometric and DXA measurements between ODM and ONDM, adjusted for age and pubertal stage

**Supplementary figure 3: Gender wise differences for metabolic measurements between ODM and ONDM**

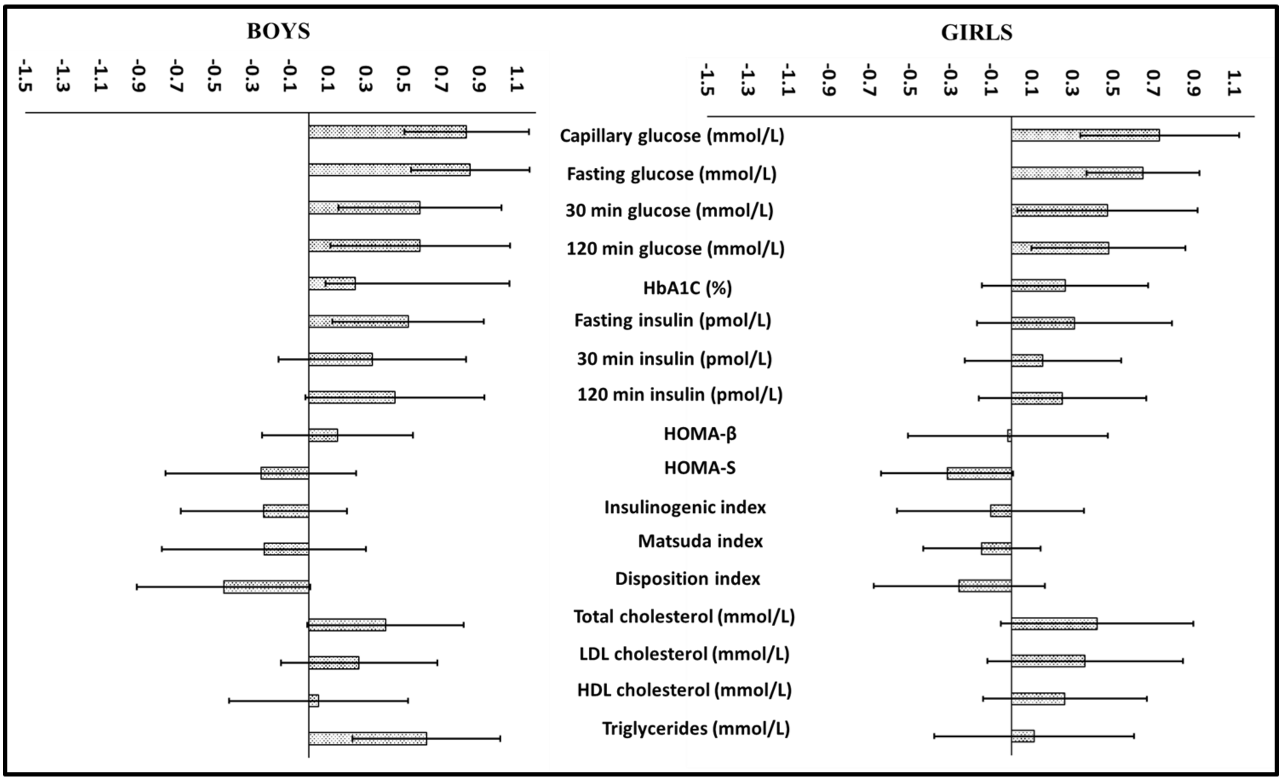


Supplementary Figure 3: Mean difference with 95% CI of SD scores for metabolic measurements between ODM and ONDM, adjusted for age and pubertal stag

**Supplementary figure 4: Overweight + obesity in offspring according to parental size in ODM and ONDM**

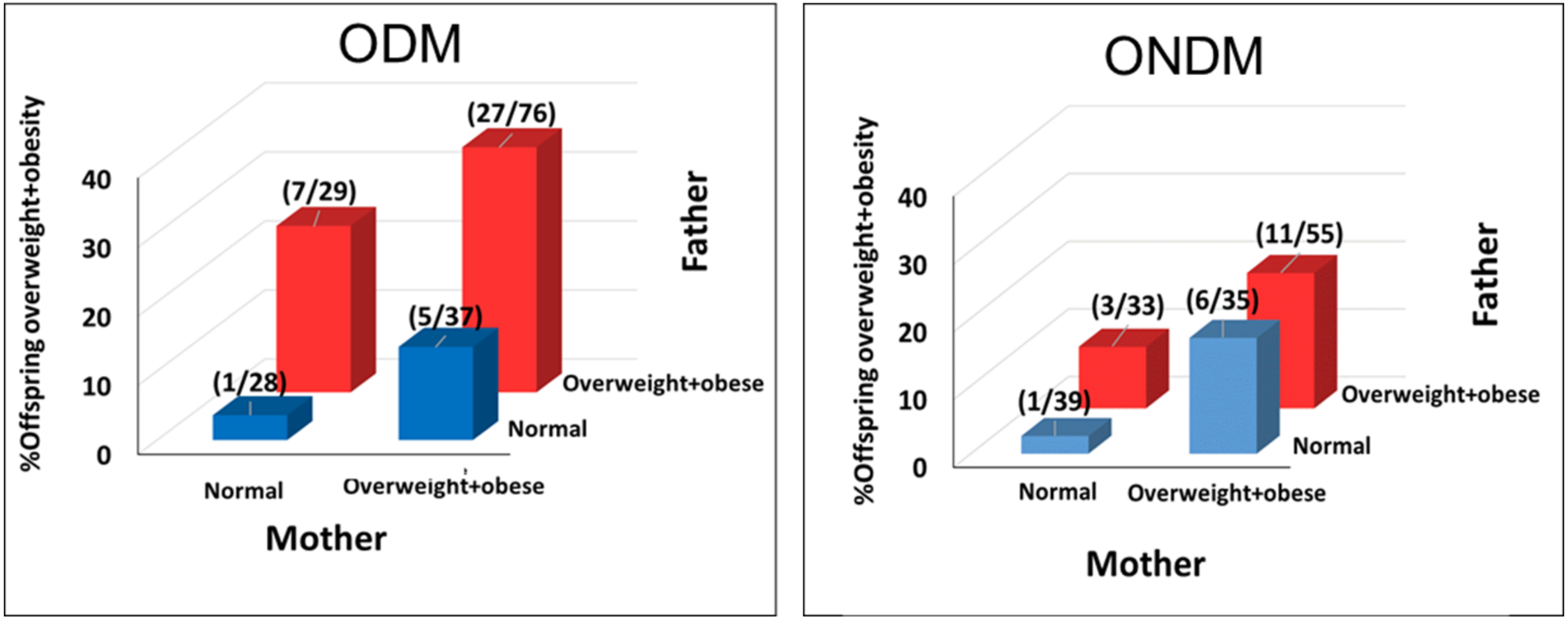


Supplementary figure 4 shows association between parental and offspring overweight-obesity, data from diabetic and non-diabetic pregnancies is combined. There is a progressive increase in the offspring overweight-obesity when none, one or both parents are overweight-obese. The association is similar whether the offspring was born in a diabetic or a non-diabetic pregnancy, though proportion of overweight-obesity was higher in children born in diabetic pregnancies. The figure highlights biparental transmission of overweight-obesity in both diabetic and non-diabetic pregnancies.

Offspring: overweight-obesity were classified using IOTF (2-18 years) and WHO criteria (>18 years and parents). Overweight-obesity in parents (WHO criteria, BMI >=25 Kg/m$^2$) was measured at follow up.

ODM: Offspring of diabetic mothers, ONDM: Offspring of non-diabetic mothers, IOTF: International Obesity Task Force, WHO: World Health Organization

**Supplementary figure 5: Agreement between venous plasma glucose and capillary glucose in parents**

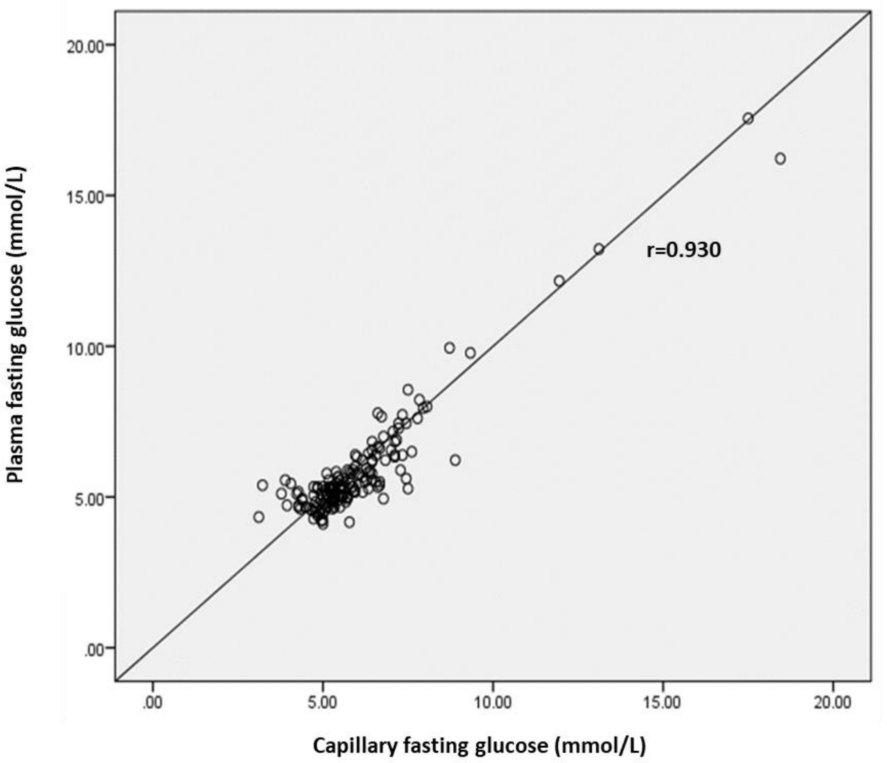


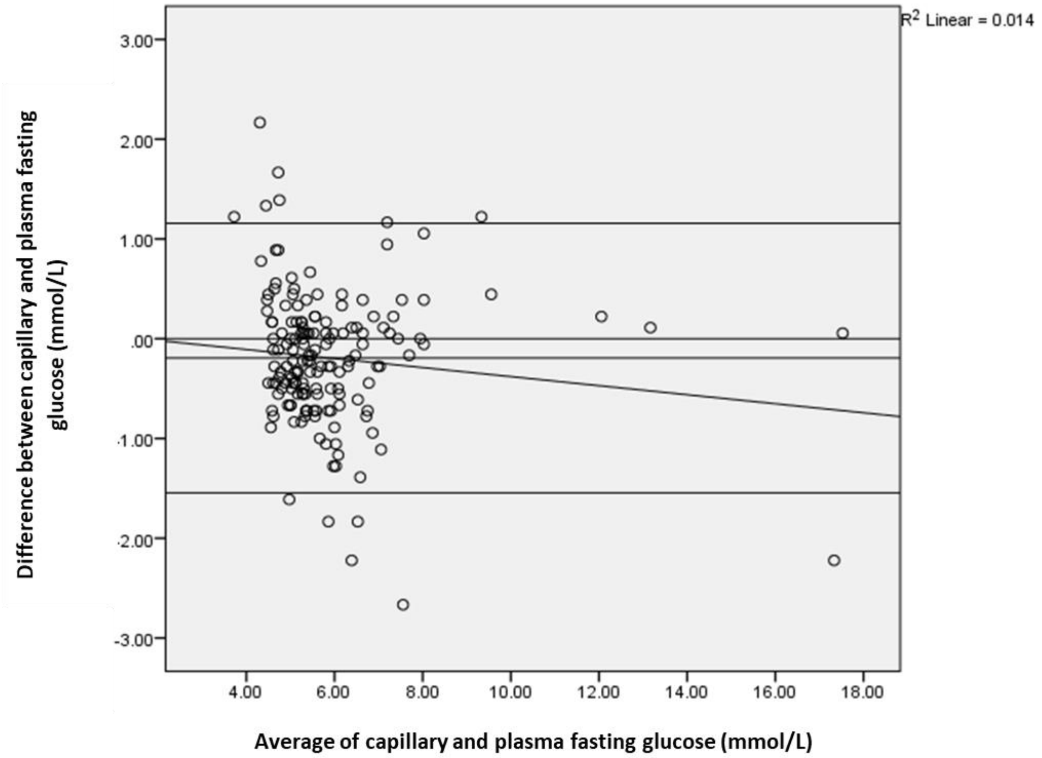


Supplementary figure:
5 a: Correlation between capillary fasting glucose and plasma fasting glucose in parents
5 b: Bland Altman Plot shows distribution of differences between venous plasma glucose and capillary blood glucose measured on glucometer vs. mean of the two measurements. A negative bias of 0.19 mmol/L is represented by line A.
